# Diurnal variation of 8-hydroxy-2’-deoxyguanosine in continuous time series of two breast cancer survivors

**DOI:** 10.1101/2023.11.18.23298714

**Authors:** Joschua Geuter, Lennart Seizer, Germaine Cornelissen Guillaume, Ayse Basak Engin, Dietmar Fuchs, Christian Schubert

## Abstract

8-hydroxy-2’deoxyguanosine (8-OHdG) is an oxidative product removed from DNA following radical oxygen species-induced damage. Being water-soluble, it can be measured non-invasively in the urine and has thus been established as a marker for ‘whole-body’ oxidative stress. Its validity and reliability as an oxidative stress marker in various chronic diseases and early carcinogenesis screening in clinical diagnosis and research are widely debated. To determine optimal measurement timing and duration, it is essential to establish the circadian profile of 8-OHdG under everyday life conditions and use reliable sampling methods. Here, we show the presence of day-night differences for 8-OHdG normalized by creatinine or urine volume in continuous time series of two breast cancer survivors who participated in integrative single-case studies and sampled their urine in 12-h-pooled collections over one month. These findings support the importance of appropriately considering the dynamic characteristics of stress indicators to reduce the risk of inconsistent or false results in clinical diagnostics.

## Introduction

Radical oxygen species (ROS) are highly reactive molecules produced either endo- or exogenously [1, 2]. They are generated during cellular metabolism and due to environmental factors such as alcoholic beverages, tobacco smoke, radiation, and stress [1–8]. If not neutralized, ROS can attack lipids, proteins, and DNA [1, 9, 10]. Under normal physiological conditions, a balance between endogenous oxidants and antioxidants exists. However, when ROS production exceeds the body’s antioxidant capacity, oxidative stress occurs [1, 2, 9–12].

One form of oxidative damage in cells involves DNA oxidation, with guanosine being particularly susceptible to such harm [4, 13, 14]. The addition of a hydroxyl group to the 8^th^ position of the guanine molecule results in the formation of 8-hydroxy-2’-deoxyguanosine (8-OHdG), a molecule initially described by Kasai and Nishimura in 1984 [15]. This oxidation of guanosine to 8-OHdG is one of the most frequent outcomes of free-radical-induced DNA damage and, consequently, one of the most extensively studied forms [9, 16].

The oxidized guanosine can pair with adenine or cytosine, potentially leading to an adenine mismatch and a subsequent transversion mutation from G:C to A:T [2, 17]. Nonetheless, repair mechanisms remove 8-OHdG from DNA, leading to its excretion from the cell [1]. Thus, 8-OHdG is considered a DNA repair product [1, 13]. Being a free deoxynucleoside, it is water-soluble and excreted into urine without secondary metabolization. Urinary 8-OHdG levels thus reflect the balance between oxidative DNA damage and DNA repair [8, 18]. Given the average rate of oxidative damage in the body, no conclusion on specific cells is possible, making 8-OHdG a marker of ‘whole-body’ oxidative damage [12, 14]. The non-invasive measurement of 8-OHdG in urine offers a significant advantage over other biomarkers, contributing to its extensive study in various health conditions, including cancer, neurodegenerative disorders, and chronic diseases [6, 10, 12, 14, 19].

To use 8-OHdG as a reliable biomarker in clinical research, understanding its excretion dynamics is essential. Existing literature presents conflicting information about whether 8-OHdG levels exhibit a circadian rhythm [4, 12, 13, 20–23]. To address this question, we analyzed urinary 8-OHdG level dynamics normalized by either creatinine concentration or urine volume in two breast cancer survivors participating in integrative single-case studies [24, 25]. This study design aims to capture the natural ebb and flow of everyday life to investigate the dynamic behavior of the variables under conditions of “life as it is lived” [26, 27]. In this evaluation of two integrative single-case studies on breast cancer survivors, time series analysis of 12-h pooled urine over 32 and 28 days was applied to reveal day-night differences in urinary 8-OHdG excretion in both patients, regardless of whether creatinine or volume correction was applied.

## Material and Methods

### Study Design

This study analyzed the urine time series from two previous integrative single case studies involving breast cancer survivors (see [24, 25]). For subject 1, the study extended over 32 days or 63 12-h intervals (from December 7th, 2004, to January 7th, 2005), while for subject 2, it spanned 28 days or 55 12-h intervals (from July 13th to August 9th, 2006). To ensure high ecological validity, minimal interference was made in the subjects’ regular daily routine. The subjects collected their entire urine, dividing it into two periods: day (from ∼08:00 to ∼20:00) and night (∼20.00 to ∼08:00). Upon collection, 0.5 g Na-metabisulfite and 0.5 g Na-ethylenediaminetetraacetic (Sigma-Aldrich, United States) were added to the polyethylene collection canisters preventing urine sedimentation and oxidation. At the end of each period, the subjects recorded the total urine volume per 12-h period, and several aliquots were directly frozen at - 20°C in 2 ml Eppendorf tubes. Once per week, samples were brought to the laboratory and stored at -70° C until further analysis. For a more detailed study design description, see [28].

Both subjects provided written informed consent to participate in the study and for data publication. The Ethics Review Committee of Hannover and Freiburg University approved the design.

### Subject Description

Subject 1, a woman in her 60s, is married with no children. She is a non-smoker and consumes moderate amounts of alcoholic beverages. Five years before the study, she was diagnosed with ductal mammary carcinoma in her left breast (pT1c, N0, M0, G2, Her2-/neu receptor-positive). The therapy included two months of radiotherapy, followed by breast-conserving surgery. One year before the study, her left breast was removed after recurrence diagnosis. Adjuvant treatment included tamoxifen and anastrozole. After therapy, she experienced cancer-related fatigue and symptoms of depression.

Subject 2, a woman in her 40s, is married and has three children. She is a non-smoker and consumes moderate amounts of alcoholic beverages. Five years before the study, she was diagnosed with ductal breast cancer in the right breast (pT2, pN1biv (6 of 13), cM0, G3, R0, ER 10%, PR 70-80%, HER2+/neu+, score = 3). Her right breast was surgically removed in 2001 (mastectomy and lymphadenectomy). Additional therapy included radiotherapy, chemotherapy according to the EC scheme (100 mg Epirubicin and 1000 mg Cyclophosphamide), and tamoxifen treatment (completed six months before the study began). After chemotherapy, she was diagnosed with secondary amenorrhea. After cancer therapy, she was diagnosed with an adjustment disorder with a depressed mood.

At the study’s outset, careful medical examinations confirmed that both subjects were clinically free of metastatic or recurrent lesions.

### Biochemical Measurements

All urinary 8-OHdG levels in the samples were analyzed in 2011 using enzyme-linked immunosorbent assays (ELISA; Kit Cayman, USA). According to the manufacturer, cross-reactivity occurs with 8-hydroxyguanosine (23%), 8-hydroxyguanine (23%), and guanosine (<0.01%). Each measurement was repeated at least twice using different aliquots and subsequently averaged. Urinary creatinine levels were determined via high-pressure liquid chromatography (Model LC 550; Varian Associates, Palo Alto, CA) [29]. All urinary aliquots from each subject were measured in a single run.

8-OHdG levels are expressed in ng/mg urinary creatinine to correct for urinary flow and glomerular filtration rate or ng of the total pooled 12-h urine (volume correction) to correct for hydration status [30].

### Statistical Analysis

All statistical analyses were conducted in *R* 4.2 [31]. Pearson correlations were calculated to assess the similarity between creatinine- and volume-corrected levels of urinary 8-OHdG. Paired, two-tailed Student’s t-tests were computed to compare urinary 8-OHdG levels during day and night periods. Additionally, autocorrelation functions (ACF) of the time series were computed to investigate serial dependencies [32], followed by least squares spectral analysis [33] and one-way analysis of variance (ANOVA). In all analyses, statistical significance was considered at p < 0.05.

## Results

The levels of creatinine- and volume-corrected 8-OHdG time series in consecutive pooled 12h-urine collections for both subjects are depicted in Figure 1. Descriptive statistics for all time series are shown in Table 1. Notably, there were no missing data. Correlations between values from the two normalization approaches, volume and creatinine correction, were significantly positive across subjects (r = 0.73, p < 0.01). In both subjects and with both normalization methods, urinary 8-OHdG levels exhibited higher variability in night samples compared to day samples. Additionally, the coefficient of variation (% CV) decreased as the collection time increased (Table 1). However, variations emerged in within-subject variability and normalization methods: volume-corrected levels showed less variability for subject 1, while creatinine-corrected values exhibited less variability for subject 2 (Table 1).

**Table 1:**
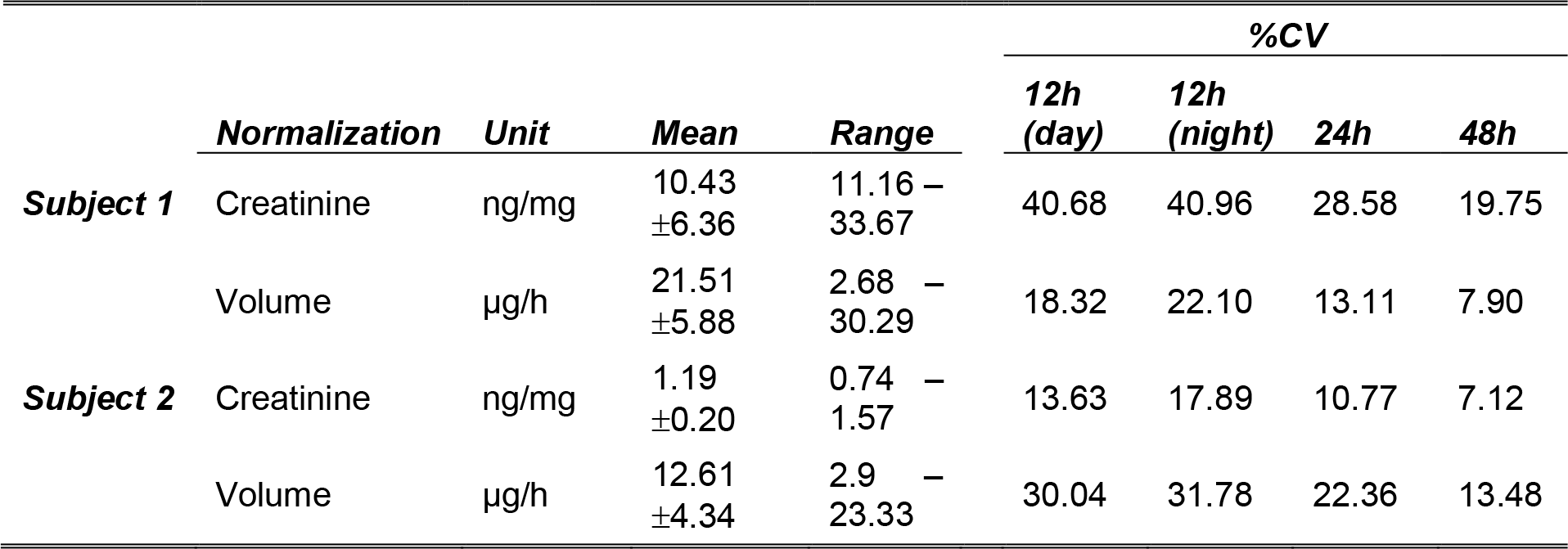
Descriptive statistics of urinary 8-OHdG. Descriptive statistics (mean, standard deviation, range) of creatinine- and volume-corrected values of urinary 8-OHdG for both subjects are shown. Coefficients of variation in percentage points (%CV) are given for 12h, 24h, and 48h collection periods.

|  | *Normalization* | *Unit* | *Mean* | *Range* | %CV |  |  |  |
| --- | --- | --- | --- | --- | --- | --- | --- | --- |
|  |  |  |  |  | *12h (day)* | *12h (night)* | *24h* | *48h* |
| **Subject 1** | Creatinine | ng/mg | 10.43<br>±6.36 | 11.16 –<br>33.67 | 40.68 | 40.96 | 28.58 | 19.75 |
|  | Volume | µg/h | 21.51<br>±5.88 | 2.68 –<br>30.29 | 18.32 | 22.10 | 13.11 | 7.90 |
| **Subject 2** | Creatinine | ng/mg | 1.19<br>±0.20 | 0.74 –<br>1.57 | 13.63 | 17.89 | 10.77 | 7.12 |
|  | Volume | µg/h | 12.61<br>±4.34 | 2.9 –<br>23.33 | 30.04 | 31.78 | 22.36 | 13.48 |

**Figure 1:**
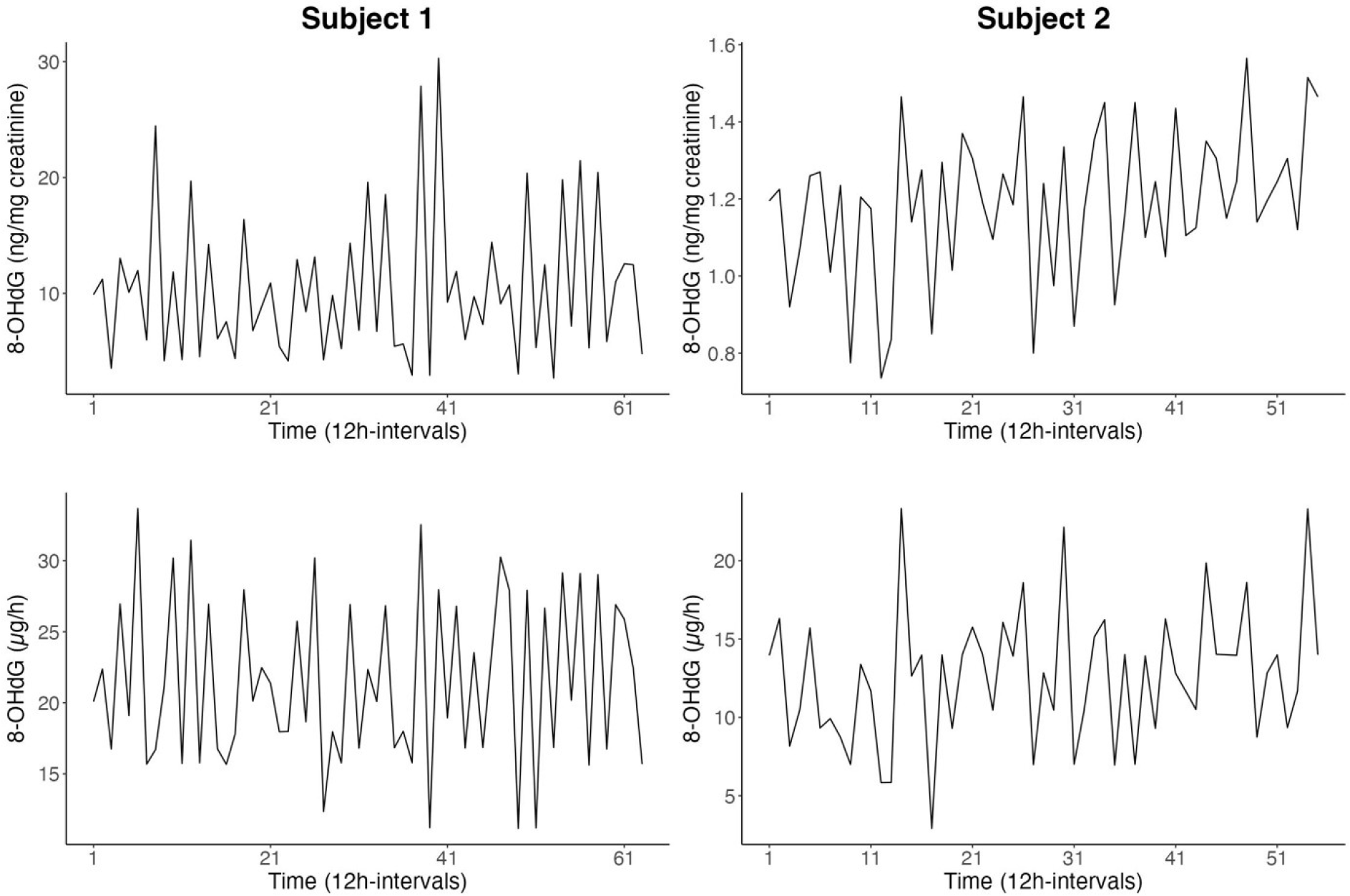
Time series of urinary 8-OHdG. The time series of urinary 8-OHdG are given as creatinine-corrected (ng/mg creatinine; top row) and volume-corrected (μg/h; bottom row) values for subject 1 (63 12h-intervals; left column) and subject 2 (55 12h-intervals; right column).

A significant difference was observed between day and night levels of urinary 8-OHdG with lower levels during the night for both subjects in creatinine-corrected (subject 1: t = -6.43, p < 0.01; subject 2: t = -2.69, p = 0.01) and volume-corrected values (subject 1: t = -7.30, p < 0.01; subject 2: t = -3.69, p < 0.01). Nevertheless, this day-night difference was not consistently expressed throughout the study period. During certain 24-h periods, an inverse relation occurred with higher values at night (Figure 2). Specifically, this applied to 6.45% of creatinine-corrected and 12.9% of volume-corrected values for subject 1 and 22.22% of creatinine-corrected and 25.93% of volume-corrected values for subject 2. Furthermore, these significant day-night differences observed for 8-OHdG normalized by creatinine and urine volume were not found for the original (uncorrected) determinations of 8-OHdG and may originate from significant day-night differences characterizing both creatinine and urine volume.

**Figure 2:**
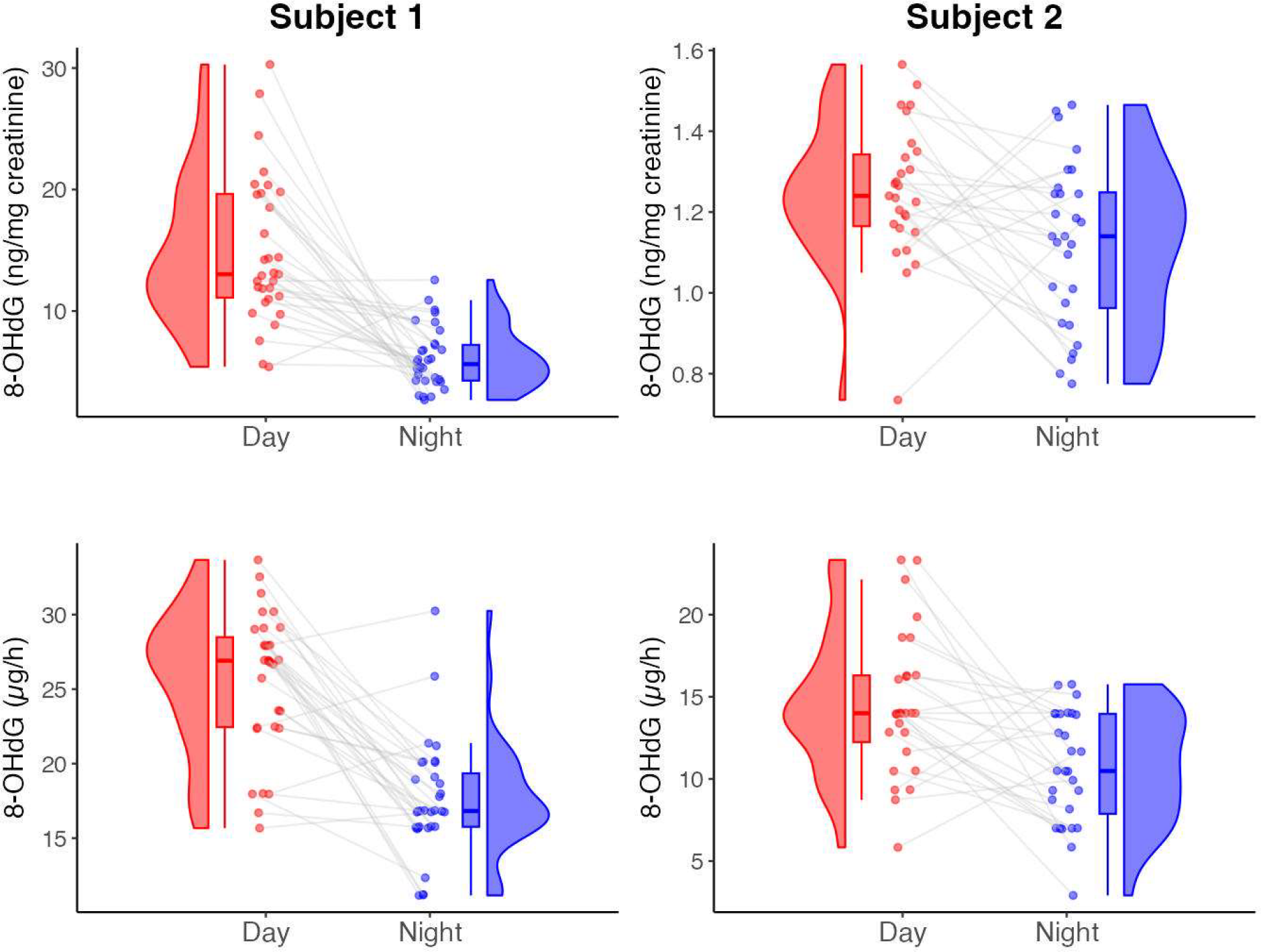
Diurnal differences in urinary 8-OHdG. Depicted is the difference between day and night values in urinary 8-OHdG as creatinine-corrected (top row) and volume-corrected (bottom row) values for subject 1 (left column) and subject 2 (right column). In each comparison, a statistically significant difference, as determined by paired, two-tailed t-tests, was found.

The autocorrelation function (ACF) plots of the uncorrected time series of 8-OHdG for subjects 1 and 2 suggest the presence of an approximately 4-day periodic component (Supplementary Figure 1). Additional analyses of variance confirm a 4-day pattern for subject 1 (*F* = 11.01, *p* < 0.01) and subject 2 (*F* = 2.78, *p* = 0.02). While a 4-day pattern is significant for creatinine (*F* = 5.59, *p* < 0.01) and urine volume (*F* = 8.84, *p* < 0.01) as well as for 8-OHdG normalized by creatinine (*F* = 10.06, *p* < 0.01) or urine volume (*F* = 8.85, *p* < 0.01) in the case of subject 1, for subject 2, only urine volume (*F* = 2.84, *p* = 0.02) and 8-OHdG normalized by urine volume (*F* = 2.83, *p* = 0.02) exhibit a 4-day pattern. An approximately 4-day periodicity is further supported by spectral analysis of uncorrected urinary 8-OHdG levels, showing a spectral peak corresponding to periods of about 90 to 121 h, accounting for approximately 50% and 21% of the total variance for subjects 1 and 2. At the common trial period of 90 h (3.75 days), phases for both subjects are similar (−282° and -276°, where 360° represents 90 h, and 0° is the start of the time series). This component (*p* < 0.05 by cosinor) has a period slightly longer than a half-week. Other variables (urine volume, creatinine, and 8-OHdG normalized by urine volume or creatinine) do not exhibit a similar spectral component that reaches statistical significance.

## Discussion

This study examined the day-night variations in normalized urinary 8-OHdG in two breast cancer survivors over a period of 63 and 55 12-h intervals (i.e., 32 and 28 days), using 12-h pooled samples. The findings strongly indicate a day-night difference in normalized urinary 8-OHdG, with significantly higher levels during the day and lower levels during the night. This rhythm was consistent across both subjects and independent of the correction method used (creatinine or volume). Importantly, this study is the first to analyze complete time series of this length, providing robust indications of significant day-night differences in corrected 8-OHdG series.

Both subjects exhibited inter-individual variations in raw and corrected 8-OHdG levels, which can be attributed to various factors such as lifestyle, diet, exposure to environmental agents, and stress [1, 10, 14, 34]. Reference values for urinary 8-OHdG levels vary across studies, with a recent meta-analysis reporting an average range of 5.9-21.6 ng 8-OhdG per mg creatinine for ELISA measurements in non-smoking subjects with a BMI <25 [10]. Notably, ELISA measurements generally overestimate 8-OhdG levels due to cross-reactivity (see Methods, [1, 22]). Although the values observed in subject 2 fall below this reference, they align with findings from other studies [11]. Given that elevated 8-OhdG levels are commonly associated with breast cancer and tend to decrease during disease progression [1, 2, 14, 35, 36], levels within the reference range might reflect a disease-free state in both subjects.

Despite the noted inter-individual differences, both subjects exhibit similar day-night variations in normalized 8-OHdG levels. The circadian rhythm of 8-OHdG has been investigated in prior studies, yielding inconsistent findings. Some studies reported circadian variation [20, 37–39], while others did not observe such a rhythm [4, 12, 21–23, 34], contrasting with our results. This discrepancy may stem from the comprehensive nature of our time series, comprising 63 and 55 continuous data points (i.e., 32 and 28 days) for the two subjects, and allowing for a more robust assessment of day-night differences. In contrast, many other studies focused on single 24-h [12, 20, 21, 23, 37, 39] or 48-h measurements [4] or sampled only once per 24-h period [34]. Our data shows that individual days may not consistently follow the typical day-night pattern, with some periods showing higher 8-OhdG levels during the night (Figure 2). Thus, an extended measurement period is essential for accurately assessing the rhythm underlying urinary 8-OHdG levels. Additionally, forced nighttime sampling in some studies [20, 37] may disrupt sleep patterns, potentially inducing psychological stress, which could influence 8-OHdG levels and dynamics [5, 6, 8, 40]. Lastly, the observed daytime peaks and nighttime throughs of 8-OHdG align with previous studies in urine and saliva [21, 23, 38, 39], though the use of 12-h urine samples limits the precise determination of peak and through timing beyond day-night differences.

When considering the day-night differences in 8-OHdG, it is important to consider the subjects’ histories of breast cancer. Although both participants were considered disease-free at the start of this study [24, 25], diseases have been shown to impact circadian rhythms [41, 42]. Given the similar rhythmicity in 8-OHdG observed in both subjects, the disease’s impact on 8-OHdG rhythmicity may have been similar, negligible, or dissipated in their current healthy state. To better understand the impact of disease on 8-OHdG rhythmicity, conducting a similar analysis in individuals without a severe disease history would provide valuable insights. Notably, subject 2’s cancer-related amenorrhea likely did not exert a discernible effect on urinary 8-OHdG levels [6].

The difference in urinary 8-OHdG levels between day and night may be attributed to increased physical and metabolic activity during the day [43] or the influence of the circadian clock on biological functions. Lee et al. [44] observed elevated 8-OHdG levels during the nocturnal period in subjects exposed to illumination before sleep, suggesting a connection to the circadian clock. Importantly, 8-OHdG as a marker of ‘whole-body’ oxidative damage, cannot provide specific insights into the cellular source of the damage [45]. Additionally, ROS can both cause DNA damage (leading to increased 8-OHdG excretion) and inhibit DNA repair (resulting in decreased 8-OHdG excretion) [45]. Without cellular insights, it is impossible to determine the cellular oxidative state solely based on urinary 8-OHdG levels. Manzella et al. [46] propose that the circadian rhythm of biomarkers of oxidative DNA damage may be influenced by the circadian activity of DNA repair systems, indicating an evolutionary adaptation to maximize protection against oxidative stress during the active phase of the day, resulting in higher excretion of 8-OHdG during that period.

Correcting raw measurements of urinary 8-OHdG is crucial to account for changes in urine concentration and prevent masking underlying dynamics [47]. As urinary samples underlie significant variations based on hydration level [48], normalization with an internal standard is essential [49], with creatinine levels or urinary volume per time commonly used as correction methods. However, both methods introduce some degree of uncertainty to the final result [50]. While creatinine correction can lead to higher uncertainty due to variations in creatinine levels independently of urinary flow, volume correction is more likely to introduce a bias towards lower values due to missed volumes [50]. As we show here, correction with either urine volume or creatinine excretion is necessary to reveal the observed day-night differences in 8-OHdG excretion, as this rhythm is not present in the uncorrected 8-OHdG time series for both subjects (Supplementary Figure 1). The uncorrected time series of 8-OHdG further show an approximately 4-day periodicity, which falls within the half-week range. This component is similar to circasemiseptan components documented for other variables such as blood pressure, heart rate, melatonin, or various cancer markers [27, 51–53]. Further study is needed to understand the significance of this periodicity in uncorrected 8-OHdG time series. Taken together, this indicates that to correctly capture the circadian rhythmicity of urinary biomarkers, it is advisable to analyze both the corrected and uncorrected time series.

Recognizing the significance of day-night differences in urinary 8-OHdG is essential for valid sampling in clinical research and diagnostics. While first-morning void samples have been correlated with 24-h pooled urine [4], they do not capture circadian variations and thus, their validity remains uncertain [4, 54]. As this study was not designed for this analysis but to investigate stress system dynamics under conditions of “life as it is lived,” it lacks a direct comparison between different sampling methods. However, the high ecological validity of the integrative single-case study approach allowed for the detection of day-night differences of 8-OHdG, highlighting the potential limitations of single measurement reductionistic study designs that often overlook broader systemic effects and interactions within the body, and take the risk of inconsistent or erroneous findings [24, 25, 55–57]. This is particularly important when using 8-OHdG as a biomarker in clinical research or diagnostics for various forms of cancer, stroke, diabetes, neurodegenerative disorders, and other oxidative damage-related chronic diseases [14, 58–60]. Sampling methods that overlook circadian rhythm may lead to false-positive or -negative results, potentially masking or exaggerating the presence of oxidative damage associated with the disease, which can lead to misdiagnosis and potentially false treatment decisions.

In conclusion, this investigation showed circadian rhythms in the urinary 8-OHdG time series of two former breast cancer patients using a research design specially tailored to the biopsychosocial characteristics of “life as it is lived” conditions [24, 25, 55, 56, 61]. Further studies in patients and healthy volunteers must now follow in order to strengthen these findings and to investigate the mechanisms underlying the link between circadian rhythms, oxidative stress and disease pathology. This should ultimately contribute to a comprehensive understanding of 8-OHdG dynamics and ROS-damage related pathologies such as cancer.

## Data Availability

All data produced in the present study are available upon reasonable request to the authors.

## Abbreviations

ROS: Radical Oxygen Species
8-OHdG: 8-hydroxy-2’-deoxyguanosine
ELISA: Enzyme-linked Immunosorbent Assay
CV: Coefficient of variation
uCr: Urinary creatinine
ACF: Auto Correlation Function

## References

1. L. L. Wu, C.-C. Chiou, P.-Y. Chang, and J. T. Wu, Urinary 8-OHdG: a marker of oxidative stress to DNA and a risk factor for cancer, atherosclerosis and diabetics, Clinica Chimica Acta 339, 1 (2004).

2. M. D. Jelic, A. D. Mandic, S. M. Maricic, and B. U. Srdjenovic, Oxidative stress and its role in cancer, Journal of Cancer Research and Therapeutics 17, 22 (2021).

3. S. Adachi, K. Kawamura, and K. Takemoto, Oxidative Damage of Nuclear DNA in Liver of Rats Exposed to Psychological Stress, Cancer Research 53, 4153 (1993).

4. M. Miwa, H. Matsumaru, Y. Akimoto, S. Naito, and H. Ochi, Quantitative determination of urinary 8-hydroxy-2′-deoxyguanosine level in healthy Japanese volunteers, BioFactors 22, 249 (2004).

5. A. Hirose, M. Terauchi, M. Akiyoshi, Y. Owa, K. Kato, and T. Kubota, Depressive symptoms are associated with oxidative stress in middle-aged women: a cross-sectional study, BioPsychoSocial Medicine 10, 12 (2016).

6. T. Iida, Y. Ito, H. Ishikawa, M. Kanazashi, R. Teradaira, A. Tatsumi, and S. Ezoe, Effects of Psychological Stress from a National License Examination on the Urine 8-Hydroxy-Deoxyguanosine Levels in Young Female Students, Taking into Account the Menstrual Cycle, Open Journal of Preventive Medicine 08, 21 (2018).

7. T. Iida, Y. Ito, M. Kanazashi, S. Murayama, T. Miyake, Y. Yoshimaru, A. Tatsumi, and S. Ezoe, Effects of Psychological and Physical Stress on Oxidative Stress, Serotonin, and Fatigue in Young Females Induced by Objective Structured Clinical Examination: Pilot Study of u-8-OHdG, u-5HT, and s-HHV-6, Int J Tryptophan Res 14, 117864692110484 (2021).

8. C. Shimanoe, M. Hara, Y. Nishida, H. Nanri, M. Horita, Y. Yamada, Y.-S. Li, H. Kasai, K. Kawai, Y. Higaki, and K. Tanaka, Perceived Stress, Depressive Symptoms, and Oxidative DNA Damage, Psychosomatic Medicine 80, 28 (2018).

9. C. N. Black, M. Bot, P. G. Scheffer, and B. W. J. H. Penninx, Sociodemographic and Lifestyle Determinants of Plasma Oxidative Stress Markers 8-OHdG and F2-Isoprostanes and Associations with Metabolic Syndrome, Oxidative Medicine and Cellular Longevity 2016, e7530820 (2016).

10. M. Graille, P. Wild, J.-J. Sauvain, M. Hemmendinger, I. Guseva Canu, and N. B. Hopf, Urinary 8-OHdG as a Biomarker for Oxidative Stress: A Systematic Literature Review and Meta-Analysis, International Journal of Molecular Sciences 21, 3743 (2020).

11. L. Chamitava, V. Garcia-Larsen, L. Cazzoletti, P. Degan, A. Pasini, V. Bellisario, A. G. Corsico, M. Nicolis, M. Olivieri, P. Pirina, M. Ferrari, M. D. Stasinopoulos, and M. E. Zanolin, Determination of adjusted reference intervals of urinary biomarkers of oxidative stress in healthy adults using GAMLSS models, PLOS ONE 13, e0206176 (2018).

12. Y.-S. Li, Y. Kawasaki, S. Watanabe, Y. Ootsuyama, H. Kasai, and K. Kawai, Diurnal and day-to-day variation of urinary oxidative stress marker 8-hydroxy-2’-deoxyguanosine, Journal of Clinical Biochemistry and Nutrition 68, 18 (2021).

13. A. Pilger, S. Ivancsits, D. Germadnik, and H. W. Rüdiger, Urinary excretion of 8-hydroxy-2′-deoxyguanosine measured by high-performance liquid chromatography with electrochemical detection, Journal of Chromatography B 778, 393 (2002).

14. E. E. M. Nour Eldin, M. Z. El-Readi, M. M. Nour Eldein, A. A. Alfalki, M. A. Althubiti, H. F. Mohamed Kamel, S. Y. Eid, H. S. Al-Amodi, and A. A. Mirza, 8-Hydroxy-2’-deoxyguanosine as a Discriminatory Biomarker for Early Detection of Breast Cancer, Clin Breast Cancer 19, e385 (2019).

15. H. Kasai, H. Hayami, Z. Yamaizumi, H. Saito, and S. Nishimura, Detection and identification of mutagens and carcinogens as their adducts with guanosine derivatives, Nucleic Acids Research 12, 2127 (1984).

16. A. Valavanidis, T. Vlachogiani, and C. Fiotakis, 8-hydroxy-2′ -deoxyguanosine (8-OHdG): A Critical Biomarker of Oxidative Stress and Carcinogenesis, Journal of Environmental Science and Health, Part C 27, 120 (2009).

17. K. C. Cheng, D. S. Cahill, H. Kasai, S. Nishimura, and L. A. Loeb, 8-Hydroxyguanine, an abundant form of oxidative DNA damage, causes G-T and A-C substitutions., Journal of Biological Chemistry 267, 166 (1992).

18. H.-W. Kuo, S.-Y. Chou, T.-W. Hu, F.-Y. Wu, and D.-J. Chen, Urinary 8-hydroxy-2′-deoxyguanosine (8-OHdG) and genetic polymorphisms in breast cancer patients, Mutation Research/Genetic Toxicology and Environmental Mutagenesis 631, 62 (2007).

19. C. Guo, P. Ding, C. Xie, C. Ye, M. Ye, C. Pan, X. Cao, S. Zhang, and S. Zheng, Potential application of the oxidative nucleic acid damage biomarkers in detection of diseases, Oncotarget 8, 75767 (2017).

20. E. L. Kanabrocki, D. Murray, R. C. Hermida, G. S. Scott, W. F. Bremner, M. D. Ryan, D. E. Ayala, J. L. H. C. Third, P. Shirazi, B. A. Nemchausky, and D. C. Hooper, Circadian variation in oxidative stress markers in healthy and type II diabetic men, Chronobiology International 19, 423 (2002).

21. R. Andreoli, P. Manini, G. D. Palma, R. Alinovi, M. Goldoni, W. M. A. Niessen, and A. Mutti, Quantitative determination of urinary 8-oxo-7,8-dihydro-2′-deoxyguanosine, 8-oxo-7,8-dihydroguanine, 8-oxo-7,8-dihydroguanosine, and their non-oxidized forms: daily concentration profile in healthy volunteers, Biomarkers 15, 221 (2010).

22. L. Barregard, P. Møller, T. Henriksen, V. Mistry, G. Koppen, P. Rossner, R. J. Sram, A. Weimann, H. E. Poulsen, R. Nataf, R. Andreoli, P. Manini, T. Marczylo, P. Lam, M. D. Evans, H. Kasai, K. Kawai, Y.-S. Li, K. Sakai, R. Singh, F. Teichert, P. B. Farmer, R. Rozalski, D. Gackowski, A. Siomek, G. T. Saez, C. Cerda, K. Broberg, C. Lindh, M. B. Hossain, S. Haghdoost, C.-W. Hu, M.-R. Chao, K.-Y. Wu, H. Orhan, N. Senduran, R. J. Smith, R. M. Santella, Y. Su, C. Cortez, S. Yeh, R. Olinski, S. Loft, and M. S. Cooke, Human and Methodological Sources of Variability in the Measurement of Urinary 8-Oxo-7,8-dihydro-2′-deoxyguanosine, Antioxidants & Redox Signaling 18, 2377 (2013).

23. I. S. Grew, V. Cejvanovic, K. Broedbaek, T. Henriksen, M. Petersen, J. T. Andersen, E. Jimenez-Solem, A. Weimann, and H. E. Poulsen, Diurnal variation of urinary markers of nucleic acid oxidation, Scandinavian Journal of Clinical and Laboratory Investigation 74, 336 (2014).

24. C. Schubert, M. Neises, K. Fritzsche, W. Geser, F. M. Ocana-Peinado, D. Fuchs, R. Hass, G. Schmid-Ott, and C. Burbaum, Preliminary Evidence on the Direction of Effects Between Day-to-Day Changes in Cellular Immune Activation, Fatigue and Mood in a Patient with Prior Breast Cancer: A Time-Series Analysis Approach, Pteridines 18, 139 (2007).

25. J. Haberkorn, C. Burbaum, K. Fritzsche, W. Geser, D. Fuchs, F. M. Ocaña-Peinado, and C. Schubert, Day-to-day cause–effect relations between cellular immune activity, fatigue and mood in a patient with prior breast cancer and current cancer-related fatigue and depression, Psychoneuroendocrinology 38, 2366 (2013).

26. L. Seizer, G. Cornélissen-Guillaume, G. K. Schiepek, E. Chamson, H. R. Bliem, and C. Schubert, About-Weekly Pattern in the Dynamic Complexity of a Healthy Subject’s Cellular Immune Activity: A Biopsychosocial Analysis, Frontiers in Psychiatry 13 (2022).

27. C. Schubert, L. Seizer, E. Chamson, P. König, N. Sepp, F. M. Ocaña-Peinado, M. Schnapka-Köpf, and D. Fuchs, Real-Life Cause-Effect Relations Between Urinary IL-6 Levels and Specific and Nonspecific Symptoms in a Patient With Mild SLE Disease Activity, Front Immunol 12, 718838 (2021).

28. C. Schubert, M. Ott, J. Hannemann, M. Singer, H. R. Bliem, K. Fritzsche, C. Burbaum, E. Chamson, and D. Fuchs, Dynamic Effects of CAM Techniques on Inflammation and Emotional States: An Integrative Single-Case Study on a Breast Cancer Survivor, Integr Cancer Ther 20, 153473542097769 (2021).

29. A. Hausen, D. Fuchs, K. Grünewald, H. Huber, K. König, and H. Wachter, Urinary neopterine as marker for haematological neoplasias, Clin Chim Acta 117, 297 (1981).

30. M. F. Boeniger, L. K. Lowry, and J. Rosenberg, Interpretation of Urine Results Used to Assess Chemical Exposure with Emphasis on Creatinine Adjustments: A Review, American Industrial Hygiene Association Journal 54, 615 (1993).

31. R Core Team, R: A language and environment for statistical computing, (2023).at <https://www.R-project.org/>

32. G. E. P. Box, G. M. Jenkins, G. C. Reinsel, and G. M. Ljung, Time Series Analysis: Forecasting and Control, John Wiley & Sons (2015).

33. G. Cornelissen, Cosinor-based rhythmometry, Theoretical Biology and Medical Modelling 11, 16 (2014).

34. M.-P. Martinez-Moral and K. Kannan, How stable is oxidative stress level? An observational study of intra- and inter-individual variability in urinary oxidative stress biomarkers of DNA, proteins, and lipids in healthy individuals, Environment International 123, 382 (2019).

35. L. M. Berstein, T. E. Poroshina, I. M. Kovalenko, and D. A. Vasilyev, Serum Levels of 8-Hydroxy-2’-Deoxyguanosine DNA in Patients with Breast Cancer and Endometrial Cancer with and without Diabetes Mellitus, Bull Exp Biol Med 161, 547 (2016).

36. F. Zahran, R. Rashed, M. Omran, H. Darwish, and A. Belal, Study on Urinary Candidate Metabolome for the Early Detection of Breast Cancer, Ind J Clin Biochem 36, 319 (2021).

37. E. L. Kanabrocki, M. D. Ryan, D. Murray, R. W. Jacobs, J. Wang, A. Hurder, N. C. Friedman, G. Siegel, B. Eladasari, B. A. Nemchausky, G. Cornelissen, and F. Halberg, Circadian variation in multiple sclerosis of oxidative stress marker of DNA damage. A potential cancer marker, Clin Ter 157, 117 (2006).

38. S. Watanabe, Y. Kawasaki, and K. Kawai, Diurnal variation of salivary oxidative stress marker 8-hydroxyguanine, Genes and Environ 41, 20 (2019).

39. D. Töbelmann and M. Dittmar, Diurnal relationship between core clock gene BMAL1, antioxidant SOD1 and oxidative RNA/DNA damage in young and older healthy women, Experimental Gerontology 151, 111422 (2021).

40. H. E. A. Çelik?, G. Tuna, D. Ceylan, and S. Küçükgöncü, A comparative meta-analysis of peripheral 8-hydroxy-2′-deoxyguanosine (8-OHdG) or 8-oxo-7,8-dihydro-2′-deoxyguanosine (8-oxo-dG) levels across mood episodes in bipolar disorder, Psychoneuroendocrinology 151, 106078 (2023).

41. S. Saha and P. Sassone-Corsi, Circadian Clock and Breast Cancer: A Molecular Link, Cell Cycle 6, 1329 (2007).

42. C. Cadenas, L. van de Sandt, K. Edlund, M. Lohr, B. Hellwig, R. Marchan, M. Schmidt, J. Rahnenführer, H. Oster, and J. G. Hengstler, Loss of circadian clock gene expression is associated with tumor progression in breast cancer, Cell Cycle 13, 3282 (2014).

43. K. Yoshihara, T. Hiramoto, T. Oka, C. Kubo, and N. Sudo, Effect of 12 weeks of yoga training on the somatization, psychological symptoms, and stress-related biomarkers of healthy women, BioPsychoSocial Med 8, 1 (2014).

44. H.-S. Lee, E. Lee, J.-H. Moon, Y. Kim, and H.-J. Lee, Circadian disruption and increase of oxidative stress in male and female volunteers after bright light exposure before bed time, Mol. Cell. Toxicol. 15, 221 (2019).

45. C. N. Black, M. Bot, P. G. Scheffer, and B. W. J. H. Penninx, Oxidative stress in major depressive and anxiety disorders, and the association with antidepressant use; results from a large adult cohort, Psychological Medicine 47, 936 (2017).

46. N. Manzella, M. Bracci, E. Strafella, S. Staffolani, V. Ciarapica, A. Copertaro, V. Rapisarda, C. Ledda, M. Amati, M. Valentino, M. Tomasetti, R. G. Stevens, and L. Santarelli, Circadian Modulation of 8-Oxoguanine DNA Damage Repair, Sci Rep 5, 13752 (2015).

47. Z. E. Melvin, H. Dhirani, C. Mitchell, T. R. B. Davenport, J. D. Blount, and A. V. Georgiev, Methodological confounds of measuring urinary oxidative stress in wild animals, Ecology and Evolution 12, e9115 (2022).

48. S. L. Nam, A. P. de la Mata, R. P. Dias, and J. J. Harynuk, Towards Standardization of Data Normalization Strategies to Improve Urinary Metabolomics Studies by GC×GC-TOFMS, Metabolites 10, 376 (2020).

49. J. Walach, P. Filzmoser, and K. Hron, Chapter Seven - Data Normalization and Scaling: Consequences for the Analysis in Omics Sciences, in Comprehensive Analytical Chemistry, Edited by J. Jaumot, C. Bedia, and R. Tauler, Elsevier (2018), pp. 165–196.

50. A. H. Garde, Å. M. Hansen, J. Kristiansen, and L. E. Knudsen, Comparison of Uncertainties Related to Standardization of Urine Samples with Volume and Creatinine Concentration, The Annals of Occupational Hygiene 48, 171 (2004).

51. G. Cornelissen and F. Halberg, Introduction to Chronobiology, (1994), p. 52.

52. F. Halberg, The week in phylogeny and ontogeny: opportunities for oncology, In Vivo 9, 269 (1995).

53. C. Schubert and C. Hagen, Bidirectional Cause–Effect Relationship Between Urinary Interleukin-6 and Mood, Irritation, and Mental Activity in a Breast Cancer Survivor, Frontiers in Neuroscience 12 (2018).

54. M. Nagao, G. Kobashi, M. Umesawa, R. Cui, K. Yamagishi, H. Imano, T. Okada, M. Kiyama, A. Kitamura, T. Sairenchi, Y. Haruyama, T. Ohira, H. Iso, and for the C. Investigators, Urinary 8-Hydroxy-2’-Deoxyguanosine Levels and Cardiovascular Disease Incidence in Japan, Journal of Atherosclerosis and Thrombosis 27, 1086 (2020).

55. C. Schubert, Soziopsychoneuroimmunologie - Integration von Dynamik und subjektiver Bedeutung in der Psychoneuroimmunologie, in Psychoneuroimmunologie und Psychotherapie, Stuttgart, Schattauer (2015), pp. 374–405.

56. C. Schubert, W. Geser, B. Noisternig, D. Fuchs, N. Welzenbach, P. König, G. Schüßler, F. M. Ocaña-Peinado, and A. Lampe, Stress system dynamics during “life as it is lived”: An integrative single-case study on a healthy woman, PLoS ONE 7 (2012).

57. L. Seizer and C. Schubert, On the Role of Psychoneuroimmunology in Oral Medicine, International Dental Journal 72, 765 (2022).

58. S. K. Urbaniak, K. Boguszewska, M. Szewczuk, J. Kazmierczak-Baranska, and B. T. Karwowski, 8-Oxo-7,8-Dihydro-2′-Deoxyguanosine (8-oxodG) and 8-Hydroxy-2′-Deoxyguanosine (8-OHdG) as a Potential Biomarker for Gestational Diabetes Mellitus (GDM) Development, Molecules 25, 202 (2020).

59. Y.-W. Hsieh, K.-C. Lin, M. Korivi, T.-H. Lee, C.-Y. Wu, and K.-Y. Wu, The reliability and predictive ability of a biomarker of oxidative DNA damage on functional outcomes after stroke rehabilitation, Int J Mol Sci 15, 6504 (2014).

60. A. Sliwinska, D. Kwiatkowski, P. Czarny, M. Toma, P. Wigner, J. Drzewoski, K. Fabianowska-Majewska, J. Szemraj, M. Maes, P. Galecki, and T. Sliwinski, The levels of 7,8-dihydrodeoxyguanosine (8-oxoG) and 8-oxoguanine DNA glycosylase 1 (OGG1) - A potential diagnostic biomarkers of Alzheimer’s disease, J Neurol Sci 368, 155 (2016).

61. L. Seizer, D. Fuchs, H. R. Bliem, and C. Schubert, Emotional states predict cellular immune system activity under conditions of life as it is lived: A multivariate time-series analysis approach, PLOS ONE 18, e0290032 (2023).

