## Supplementary Figure 1 for "Diurnal variation of 8-hydroxy-2’-deoxyguanosine in continuous time series of two breast cancer survivors"

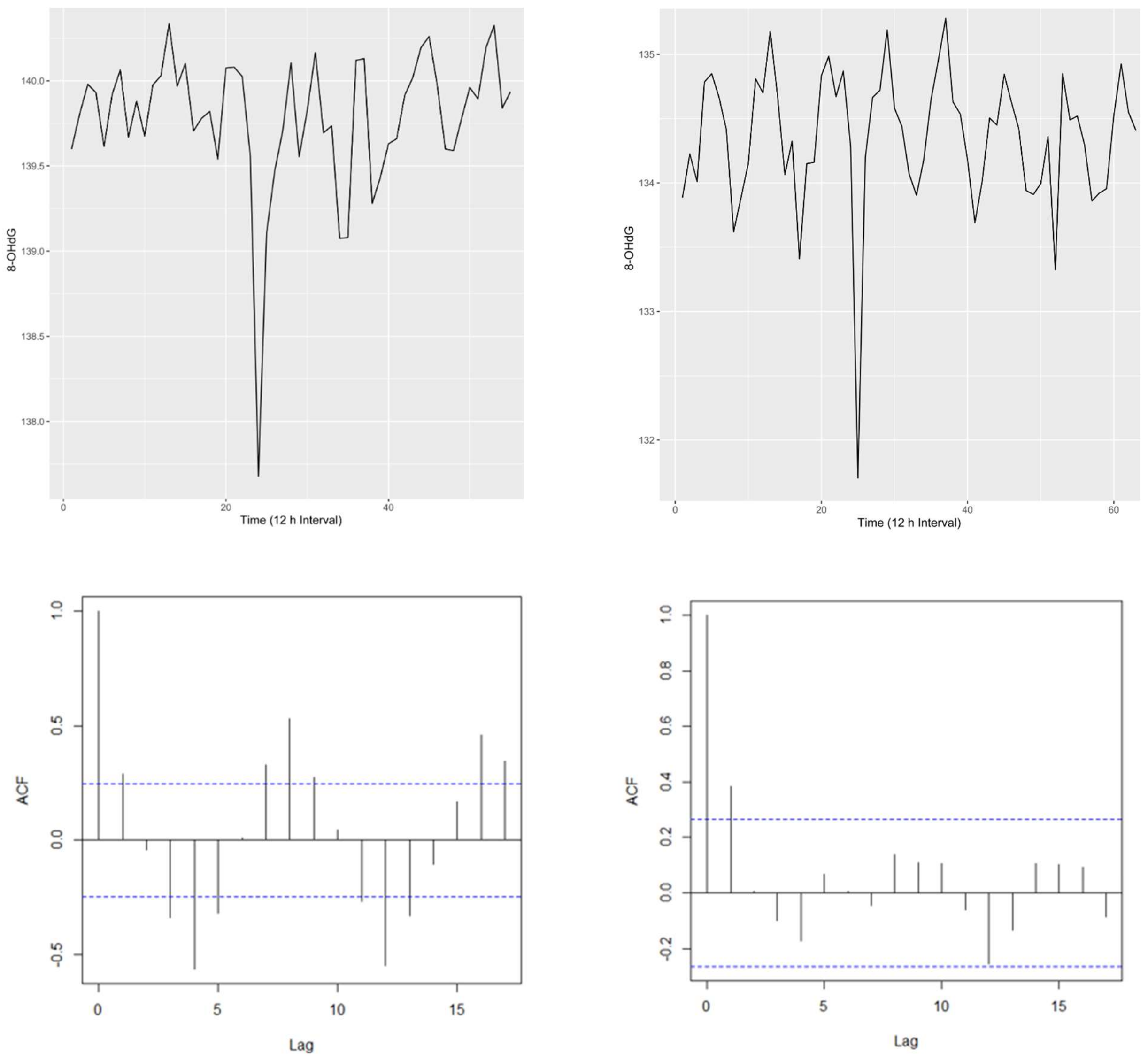


Supplementary Figure 1. Uncorrected time series of 8-OHdG (top row) for subjects 1 (left) and 2 (right) with corresponding ACF plots (bottom row).
